# Magnetic bead-based separation of functionally competent human sperms with hyaluronic acid and its superiority over conventional swim-up and density gradient methods

**DOI:** 10.1101/2025.11.10.25339963

**Authors:** Manjunath Siddaramaiah, Christy Rosaline Nirmal, Vasan S. S, Dhananjaya Dendukuri

## Abstract

The selection of competent human sperm influences the successful outcome of Assisted Reproductive Technology. Conventionally used techniques like density gradient and swim-up, use motility-based separation, which may not correlate with the maturity and quality of the sperm. Existing techniques that select mature sperm based on hyaluronic acid binding ability, select very small numbers of sperm, rendering them unsuitable for intra-uterine insemination. We report the development of a new method, USelect that selects and isolates millions of mature sperms based on their ability to bind to hyaluronic acid. Validation of Uselect was done on semen samples (n=40) compared with the conventional methods, like density gradient, and swim up. Subsequently, Uselect selected sperms were used for intra-uterine insemination in a clinical trial (n=50 female patients). Sperm isolated using USelect showed similar mean sperm count and rapid progressive motility compared to SU, but lower than the density gradient method. However, the DNA fragmentation index and hyaluronic acid binding capacity were statistically significant in the USelect sperms that promoted the fertilization rate and showed a higher percentage of biochemical pregnancy (32%, n=16) than density gradient (18%, n=9).

## INTRODUCTION

Infertility is a global issue impacting multitudes of people ranging between 48.5 million to 186 million, leading to notable repercussions such as societal stigma and financial challenges [1,2]. Approximately 15% to 20% of couples seeking medical help with conception are affected by infertility [3]. In 2022, Cox *et al.* found high infertility rates globally and regionally, emphasizing the need for improved data collection to enhance fertility care services and support those affected [4]. Ranging from the simple IUI to the more complex Intra Cytoplasmic Sperm Injection (ICSI) technique various Assisted Reproduction Technology (ART) options are available, that vary in efficiency based on factors such as sperm quality, and female factors. Novel sperm isolation methods do not reduce the importance of natural selection barriers around the oocyte. Rather, they are aimed at increasing the likelihood of a fertilization event by enhancing and refining the selection process, particularly in cases where the sperm may have difficulties in naturally navigating these barriers [5,6]. The goal remains to enhance the success of intrauterine insemination (IUI) cycles while acknowledging the crucial role that natural selection plays in the process of fertilization [7]. IUI is the simplest ART method, it can be conducted using the natural cycle and is currently accessible in various countries. It involves the direct administration of prepared sperm into the uterus using a catheter and does not demand any specialized equipment [8]. However, with a success rate of 7-14%, IUI is significantly less successful than IVF and ICSI [9]. On the other hand, both IVF and ICSI are resource-intensive, require non-natural fertilization, and are financially challenging for many couples [9]. Therefore, there is a need for methods to improve the success rate of IUI and provide higher rates of fertilization.

Around 40% to 50% of infertility cases are due to male infertility, with abnormal semen [10]. The decline in sperm quality and concentrations can be attributed to various environmental and lifestyle factors [11], highlighting the importance of sperm selection before applying ART. Traditionally, semen analysis has focused on sperm count, motility, and morphology, although they are not specific indicators of sperm functionality. In some cases, morphologically abnormal sperm may be viable, while sperm that appear “normal” may not be [12]. Additionally, immotile sperm may have better, more mature DNA content than motile sperm [13]. Swim-up (SU) is the most used technique for sperm separation, where sperm are separated based on their increased motility and the ability to swim out of seminal plasma [14]. Density gradient (DG) centrifugation is the preferred technique for severe oligozoospermia, teratozoospermia, or asthenozoospermia cases to separate a greater number of motile spermatozoa from seminal plasma [15]. However, these two techniques rely solely on the motility of sperm, which is important in natural conception and IUI until the sperm reaches the oocyte. Utilizing electric fields, the Zeta method and the Cell Sorter-10 achieve electrostatic charge-driven sperm separation, focusing DNA integrity for isolating mature sperm. However, limitations include varying percentages of sperm selection, the need for specialized equipment, and technical expertise for precise implementation [16]. Techniques such as magnetic activated cell sorting (MACS) use negative selection procedures to remove non-viable sperm and do not specifically select based on their ability to fertilize an egg. The functional properties of sperm, such as acrosomal status and DNA maturity, become important factors once the sperm reaches the fallopian tube for successful fertilization of the oocyte [17,18]. The ability of human sperm to bind to hyaluronic acid (HA) indicates their DNA maturity, viability, and unreacted status of the acrosome [19]. Studies have shown that HA-selected sperm results in improved clinical pregnancy rates compared to conventionally isolated sperm [20,21].

Due to its benefits, HA selection has become a popular method for sperm preparation in ART procedures. Currently, two widely used sperm-HA binding selection systems are Physiological Intra-Cytoplasmic Sperm Injection (PICSI) and SpermSlow. However, these techniques have certain procedural drawbacks. PICSI is not suitable for selection and isolation due to uneven hydration or detachment of the entire HA microdot from the surface. SpermSlow, on the other hand, is a viscous media mainly consisting of HA and requires careful handling of the delicate and flexible HA microdot or microdrop. Additionally, these techniques cannot be used for IUI as they only select a few HA-binding sperms, and their high cost and low yield of mature sperms. To address these issues, a novel magnetic bead-based sperm separation method called USelect has been developed, which utilizes the HA-binding method to select millions of functionally competent human sperm, enabling faster processing in ART.

## MATERIALS AND METHODS

### Ethical Approval

As per the relevant guidelines and regulations, all methods employed in this study were conducted following the approved protocol (Protocol No. MAARS-01/2015, dated 14 Nov 2015) and followed the standards set forth by the Independent Ethics Committee, as per Central Drugs Standard Control Organization (CDSCO) regulations. Additionally, the study has been registered in the Clinical Trial Registry of India (CTRI, Trial acknowledgment number REF/2015/09/009843) to ensure transparency and adherence to ethical and regulatory standards.

### Statistical Analysis for Sperm Separated by conventional and USelect Techniques

The sample size was calculated based on pilot experimental data. At a typical statistical significance level of 5% (α = 0.05), a widely accepted power of the trial to be 0.8 (80%), and using an effect size of 0.6 for the control and the experimental (USelect) groups, the sample size required per group was calculated to be 44. All statistical analyses were performed using IBM SPSS Statistics ver. 20.0 (IBM Corp, USA). Descriptive statistics for each outcome variable including mean, median, SD, range, etc. were evaluated for each separation technique group as well as the native sample. Comparison of means of all the outcome variables, i.e., motility (% progressive motility), morphology (% normal forms), and DNA integrity (DFI) was performed using Student’s t-test and one-way ANOVA with post-hoc Tukey tests, at 95% CI (p < 0.05 taken to be statistically significant).

### Inclusion exclusion criteria

The study participants included in the study were limited to the male partners of infertile couples who were referred for semen analysis to the Andrology Laboratory. Additional inclusion criteria required that subjects must be 18-45 years old with normal health as per medical history and routine physical examinations, and not shown positive for pregnancy test for at least one year. Men aged >45 years, diagnosed with diabetes or other serious, long-term illnesses, those who tested positive for HIV, HBV, HCV, etc., and patients who may not be able to provide semen samples, based on their medical history and physical examination were excluded from the study. All the subjects included in the study signed informed consent and provided a semen sample by masturbation after 2-4 days of abstinence.

### Immobilization of HA on Magnetic Beads

The biotinylated hyaluronic acid polymer, 10mg/mL (10 kDa, Creative Pegworks) in 1X PBS buffer was mixed 1:1 ratio with 10mg/mL of streptavidin-coated magnetic beads (MyOne Streptavidin C1 Dynabeads, Invitrogen). The mixture was then incubated on a rotary mixer for 1 hour at room temperature. The hyaluronic acid (HA)-coated beads were kept on the DynaMag magnetic stand (Invitrogen) for 2-3 minutes and the supernatant was separated. The beads were washed 3 times using 1X PBS containing 0.05% Tween-20 and 0.1% BSA and re-suspended in 100 μl of the same buffer. The immobilization was confirmed by flow cytometry assay [29] using FITC conjugated anti-HA antibody. The binding of the antibody to the HA on the beads generates a measurable fluorescent signal, which is then detected and quantitatively analysed by a flow cytometer, providing insights into the efficiency of the immobilization process. This method ensures the reliable coating of magnetic beads with HA and offers a quantitative means of validation through fluorescence-based analysis.

### Acute in vivo toxicity assay

An acute *in vivo* toxicity assay was conducted to validate the abnormal toxicity assessment of Uselect. The study, performed by Bioneeds India Private Limited in Bangalore, utilized Swiss albino mice. The male mice, aged 8 weeks and weighing between 19.23 and 21.56 grams, were divided into two groups, each comprising 5 animals. The test substance was administered intravenously through the lateral tail vein, with a dosage of 0.5 mL per animal. The concentration of the test substance was 10 mg/mL, with a bead concentration ranging from 6 to 12 * 10^9 beads/mL.

Throughout the 48-hour observation period following administration, a meticulous monitoring approach was employed to promptly detect any potential anomalies or unfavorable reactions in the mice. This experimental setup adheres to the standard procedures for acute in vivo toxicity assessment and ensures a comprehensive evaluation of Uselect’s safety profile in the specific context of intravenous administration in Swiss albino mice.

### Isolation of HA Binding Sperms

The sperm samples used in this study were either collected and stored in liquid nitrogen or freshly collected from the study participants. Liquid nitrogen stored samples were thawed by placing the vial in a 37°C water bath for 10-15 minutes. The liquefaction and sample preparation were carried out as described previously [30]. For initial optimization steps, 6 μl of HA-coated magnetic beads (10 mg/ml) was added with 0.1 ml of the semen sample. For clinical validation studies, the test volume was increased with 24 µl of HA beads for 1 ml of semen sample. Briefly, to the immobilized HA-coated beads 0.1-1 ml of pre-processed semen sample was added and incubated for 15 minutes at 37°C. The tube was then kept on the magnetic stand to separate unbound sperm from HA-bead-bound sperm. The unbound sperms in the supernatant were separated using a pipette. Bead-bound sperms were then suspended in 100 µl of Sperm Wash Media (SWM) containing 100 units of hyaluronidase enzyme (Hyaluronidase from bovine testes-Type IV-S, Sigma-Aldrich) and incubated for 15 minutes at 37°C. The vial was placed on the magnetic stand and the supernatant, which contains the sperms was collected for motility and sperm count analysis. The effect of HA-bead concentration on sperm binding, hyaluronidase enzyme concentration, incubation time, and pH on sperm release was studied.

### Semen sample preparation using conventional swim-up and density gradient methods

As described previously [14] the semen samples used in this study were prepared using conventional (SU and DG) methods by trained embryologists, and the comparative analysis was carried out in comparison to the novel USelect technique. Briefly, the sample was prepared for SU by adding 0.5 mL of semen sample with 1 mL of Sperm-Wash media (FertiPro NV, Belgium) and centrifuged for 10 min at 1,200 rpm. The supernatant was discarded, and sperm wash media was slowly layered over the pellet along the sides of the tube. To increase the surface area the tubes were kept at an angle of 45° in the incubator at 37°C for 45 min. After incubation, the tubes were returned to the vertical position and 1 ml of the supernatant was gently removed, aspirating the sperm from the upper meniscus downwards with a sterile pipette.

In the DG method, a two-layer density gradient formed by an upper layer of 40% (v/v) and a lower layer of 80% (v/v) media (FertiPro NV, Belgium) was incubated at 37°C. On top of the 40%-layer, 0.5 mL of semen sample was layered without disturbing the interface. The tube was centrifuged for 15 minutes at 1200 rpm. Motile spermatozoa migrate through the distance between the layers and are concentrated in the lower layer (80%). After centrifugation, the supernatant was gently removed, and the pellet was re-suspended in 2 ml of HEPES-based Sperm-Wash media (Sure Life, Singapore) and centrifuged at 1200 rpm for 7 min to remove traces of DG media. The supernatant was discarded, and the pellet was re-suspended in Sperm-Wash media. The samples prepared by both methods were used for the determination of motility, morphology, and DFI.

### Comparative validation of functional parameters in USelect, DG, and SU

The functional parameters of the sperm prepared using USelect were compared with the conventional sperm separation techniques. This is a multi-arm study that has SU, DG, USelect techniques, and unprocessed (native) samples, to compare the functional parameters of motility, morphology, and DNA integrity of sperms separated by these techniques. This 6-month study was conducted at Manipal-Ankur Andrology and Reproductive Services, Bangalore, India.

Forty-five male partners from infertile couples, who met the inclusion/exclusion and additional sample criteria, were recruited into the USelect, DG, and SU comparison study. However, due to patient dropouts, experimental inadequacy, outliers, etc., the final number for analysis was reduced to 40. The mean ± SD age of the subjects was 34.00 ± 4.00 years. One outlier sample with unreasonably high DFI after processing by the three methods was excluded from the analysis. For the sample prepared by USelect, SU, and DG the routine semen analysis such as sperm count, motility, and morphology were performed according to WHO 2010 guidelines^31^. DNA Integrity of sperm in the native, unprocessed sample was determined using flow cytometry. Following sperm separation techniques, namely SU, DG, and the novel USelect separation techniques the sperm characteristics were determined and compared with the pre-processed native sample. The Rapid progressive motility of the sperms was defined as >25 μm/s, at 37°C, and was determined according to WHO 2010 guidelines for the native and post-process sperms, to compare the separation techniques. The percentage of normal and abnormal forms of sperms in native, as well as post-processed samples, were determined according to WHO 2010 guidelines [31].

### DNA Integrity

Flow cytometry (FCM) was used to determine the DNA integrity in terms of DFI for pre and post-processed samples as described by Evenson *et al.,* with minor modifications [32] with FITC conjugated anti-HA antibody. The DFI for the samples was calculated using the percentage of viable sperms in the sample as described previously by the hypo-osmotic swelling test (HOS). Samples that have DFI ≤ 15% were considered to have ‘good’ DNA integrity followed by DFI 15% - 30% as ‘moderate’ and DFI > 30% as ‘poor’ quality sperms.

### Clinical Validation by Comparing USelect and Density Gradient Methods

The study was extended by two years to validate the method through a clinical trial where the USelect method was used to isolate viable sperm for use in IUIs conducted on human patients. In this trial, biochemical pregnancies achieved using the USelect method were compared with those achieved using the conventional DG method. The inclusion and exclusion criteria used for the trial were the same as the comparative study. The extension study was conducted by the stratified, randomized selection of patients concerning the time of enrolment to the study. The first 50 women enrolled for the study were considered for the Uselect group and the next 50 for the DG group. A sample was collected from the male partner of the patient on the day of the IUI procedure along with the informed consent form. Processed samples (Uselect and DG) were handed over to the clinician to perform the IUI. *Beta* Human c+horionic gonadotropin (®-hCG) levels were estimated at the end of three weeks followed by an ultrasound scan taken at the end of the sixth week to confirm the clinical pregnancy. Four patients under the USelect group and nine patients under the DG group were considered for a second IUI cycle. The decision to take the patient into the second IUI cycle under each group was made solely by the treating clinicians based on the clinical findings and laboratory investigations.

## RESULTS

To isolate sperm with the ability to bind to HA, HA-conjugated magnetic beads were prepared in-house by conjugating streptavidin-coated silica-based magnetic beads with biotinylated HA. The immobilization of the HA biotin with the magnetic beads was further confirmed with a flow cytometric assay. The resulting HA-functionalized magnetic beads were then used to capture HA-bound sperm from semen samples.

In brief, semen samples with HA-competent and HA-non-competent sperm were incubated with HA-coated magnetic beads. HA-competent sperm (light green in Figure 1) selectively bound to the beads, while non-competent sperm remained unbound. A magnetic field separated bead-bound from unbound sperm, and the latter was washed away. To release the bead-bound sperm, the HA-bound sperm were treated with the naturally occurring enzyme hyaluronidase, which cleaves the HA molecules and frees the sperm. The released sperm were then separated from the magnetic beads using a magnetic field, allowing for ART usage. The schematic representation in Figure 1 provides an overview of all the steps involved in the protocol. This approach offers a specific and efficient method for isolating HA-bound sperm, which may be useful for studying sperm function and identifying subpopulations of sperm with specific fertilization potential.

**Figure 1:**
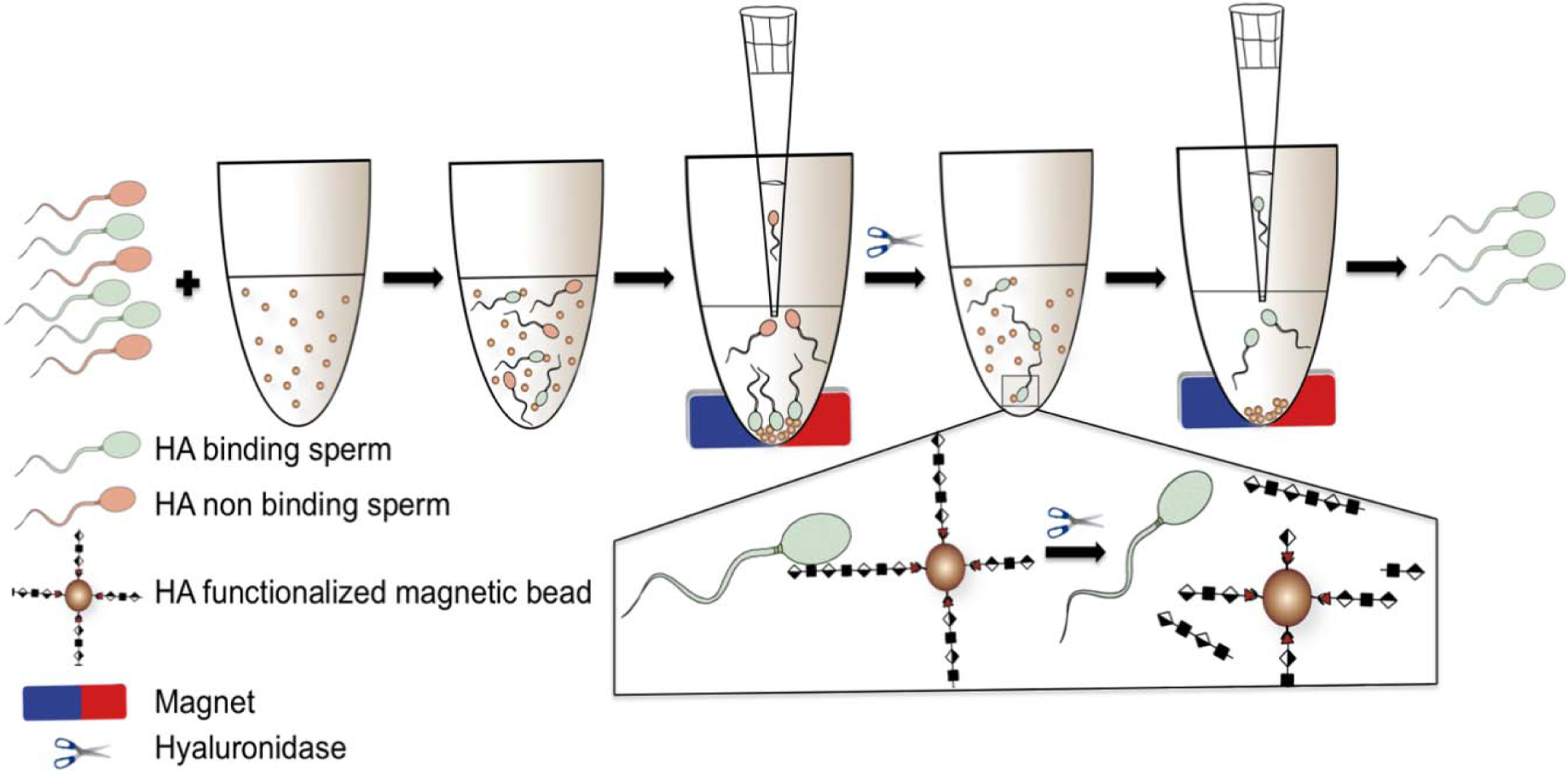
Schematic Overview of HA-Bound Sperm Isolation Protocol. This figure illustrates the step-by-step procedure for isolating HA-bound sperm. Semen samples containing HA-competent (light green) and HA-noncompetent (brown) sperm are incubated with HA-functionalized magnetic beads. The competent sperm bind to the beads, while non-competent sperm remain unbound. A magnetic field separates bead-bound sperm, which are retained, from unbound sperm, which are washed away. Hyaluronidase is used to release bead-bound sperm by cleaving HA molecules, followed by magnetic field separation to isolate the released sperm for assisted reproductive technologies. This method provides precise isolation of HA-bound sperm for the study of sperm function and identification of specific fertilization potential subpopulations.

Using this method, a significant majority of the magnetic beads were effectively removed before transferring the isolated sperm to the patient, leading to the exclusive collection of HA-competent sperms in the newly prepared tubes. The possibility of toxicity by any remnant beads post-IUI was assessed on mice models. The animal’s initial body weights were measured on the study’s first day, establishing a baseline against which any weight changes during the observation period could be evaluated. Throughout the entire 48-hour post-intra venous administration phase of the test item, none of the animals displayed clinical signs indicative of toxicity. Notably, there were no instances of recorded mortality within this time frame. This collective evidence strongly suggests that Uselect did not result in noticeable adverse effects or fatalities within the stipulated timeframe.

To optimize the protocol, three key variables – 1) the mass of HA-coated magnetic beads, 2) the amount of enzyme used and 3) the pH of the buffer containing the enzyme were individually optimized. The percentage of bound sperms increased from 40% to 60% with an increase in HA-bead concentration from 20 to 60 μg respectively; however, the increased concentration of beads beyond 60 ug shows reduced binding of the sperms. Three different concentrations of Hyaluronidase, 10, 50, and 100 U, were tested with 100 μl of sperm samples bound to HA beads, by incubation at 37°C for 15 minutes. The sperm release was increased by 100% and 116% at 50 and 100 U of an enzyme, respectively when compared to the results using 10 U of an enzyme. When using 100 U Hyaluronidase enzymes, 2.6 million sperm were obtained from 100 μL of semen sample. Both neutral and acidic pH (7. 0 and 5.0) were tested to study the effect of pH on the Hyaluronidase-based release of HA-bound sperm (Supplementary Fig. S1).

It was observed that both motile and immotile sperms were able to bind to HA beads. The number of sperm bound to HA beads varied between samples, resulting in 8-46% (average 27.6%) bound sperms and 54-92% (average 72.4%) unbound sperms (Figure 2).

**Figure 2:**
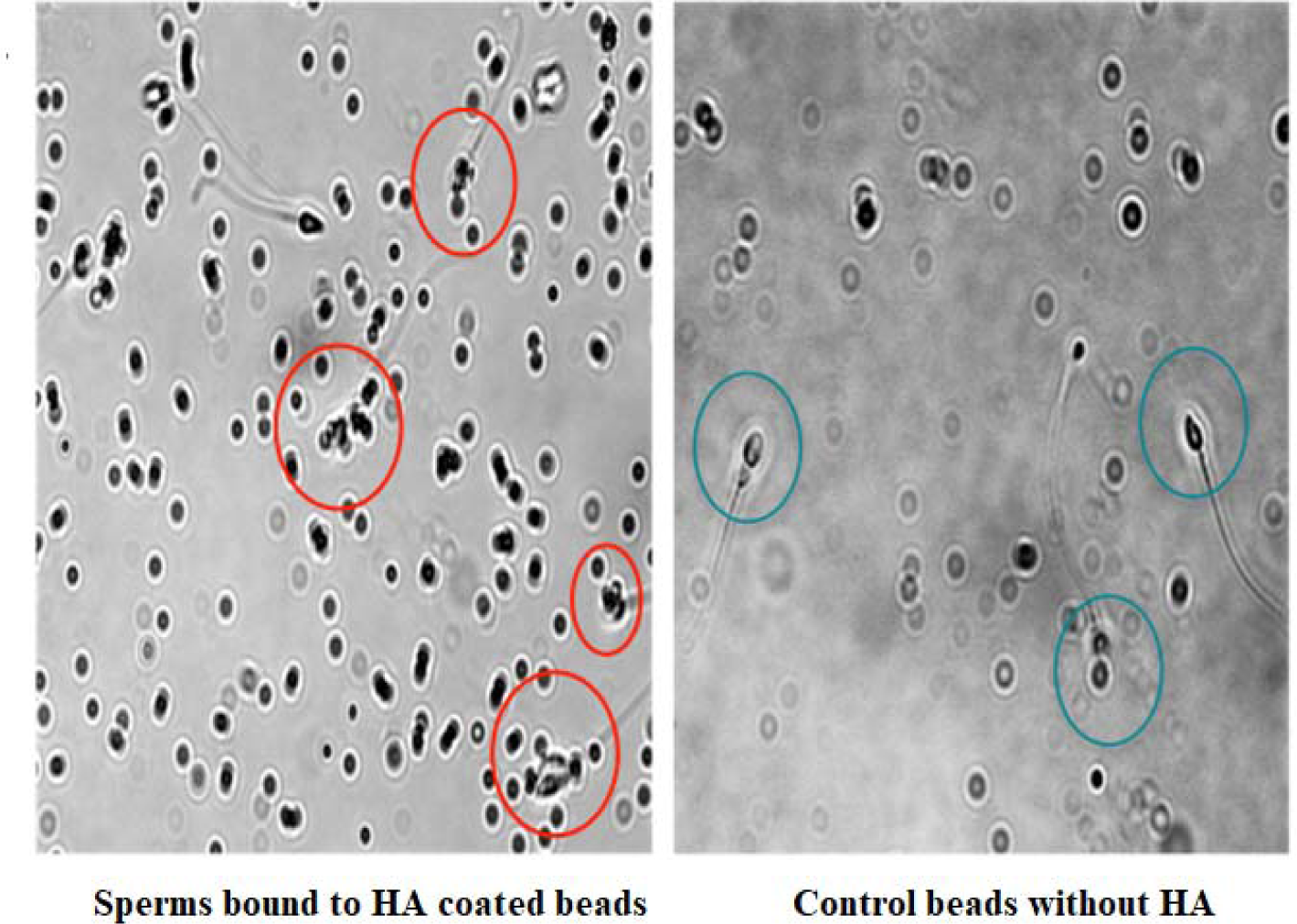
Isolation of HA binding sperms using HA functionalized magnetic beads.

The motility of the sperms obtained after HA binding was dependent on the sample, with a range of 28-58% motility of its initial sample, but after being treated with Uselect, the motility increased to 60-90% (Figure 3). The sperm count was reduced after processing the semen samples with Uselect, as the magnetic bead-based method selectively isolates only highly HA-competent sperm (Figure 4).

**Figure 3:**
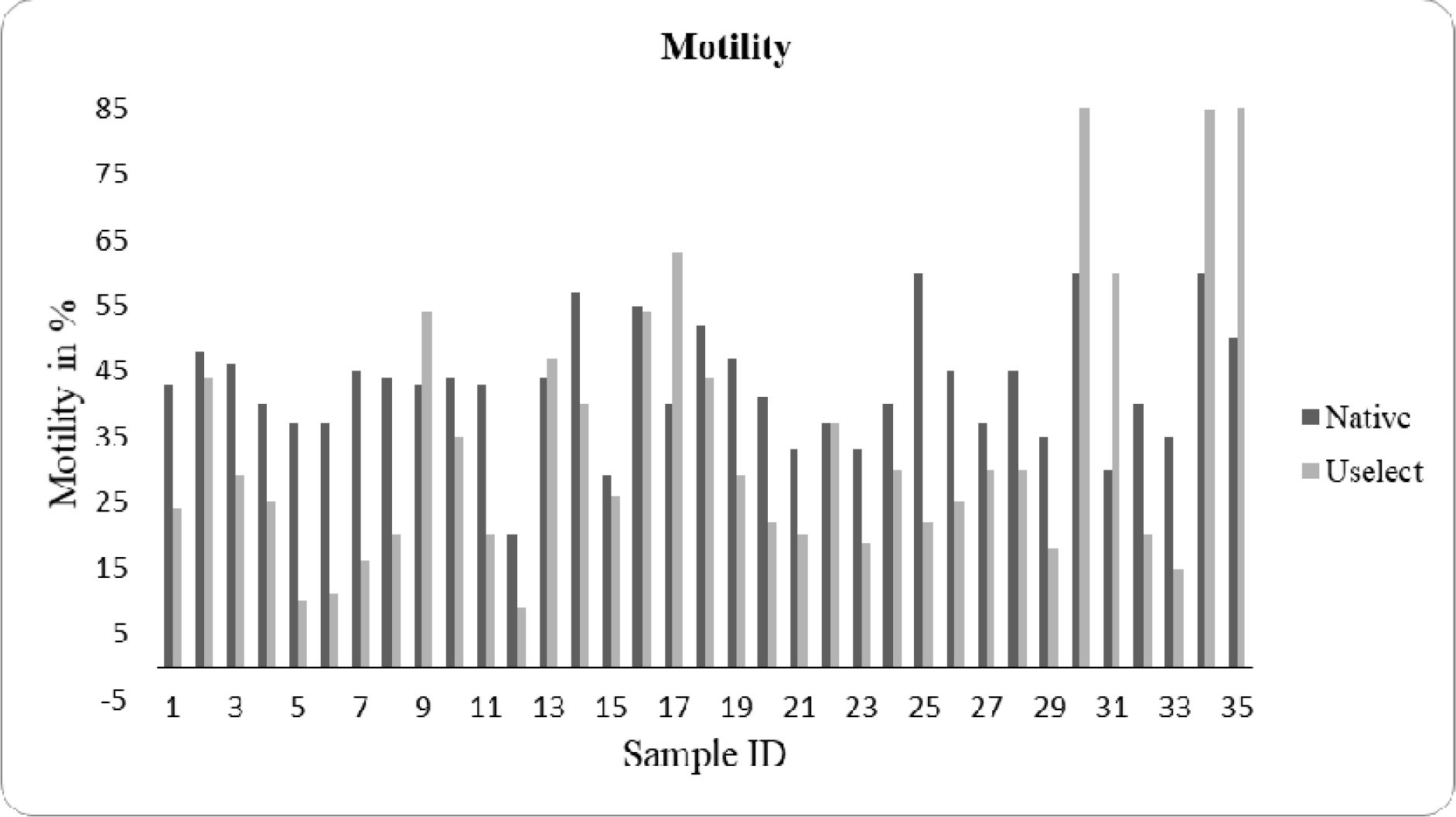
Comparison of motility of initial and HA bead isolated sperms.

**Figure 4:**
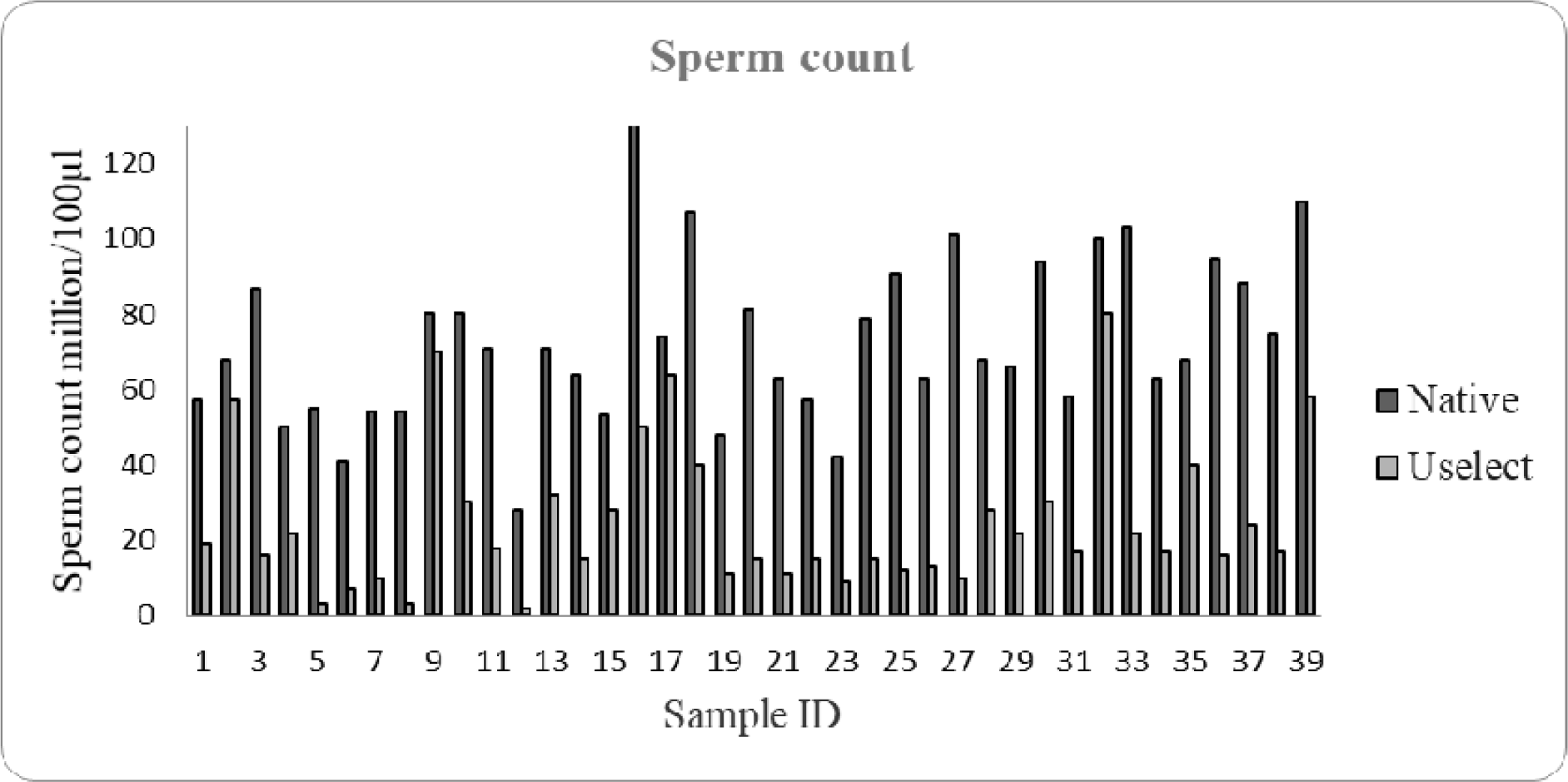
The estimated population of sperms obtained after HA binding and enzyme treatment.

The optimized USelect technique was further evaluated against SU and DG methods.

In the initial phase of the study, a total of 45 male partners who were part of infertile couples were enlisted as participants. However, as the study progressed, 5 individuals voluntarily withdrew themselves from the study. Additionally, one participant’s sample was excluded from the analysis due to an insufficient volume of sample provided. This exclusion was based on the criteria of sample adequacy set for the study.

The mean ± SD and other descriptive statistics of routine semen analysis parameters and DNA Fragmentation Index (DFI) of unprocessed samples and sperms separated using USelect, DG, and SU techniques are shown in Table 1. Post-processing, the mean sperm count was significantly different for the three separation techniques (P <0.001) in comparison with the native unprocessed sample. The sperm count for USelect was like SU, while the motility was lower than the other techniques. Morphology after the USelect separation process, in terms of the percentage of normal forms, is maintained in the same range and was not significantly different (P = 0.99) from that of the native group. The proportion of sperms with poor DNA integrity was found to be significantly reduced in the sperms selected using the USelect separation process as compared to the native sample. Figure 5a shows the flow cytometry output gated to differentiate the debris (extreme left), DNA fragmented sperm population at the bottom side of the chart, and cells with the un-fragmented DNA at the upper part of the chart. Figure 5b summarizes the variation in DFI of the sperms isolated using USelect, DG, and SU techniques. To study the improvement in DNA integrity of the sperms separated by USelect, the subject population was classified into three sub-groups based on the DFI of native samples. USelect could pick up sperms with good quality DFI from the ‘moderate’ and ‘poor’ DFI subgroups. No further enhancement in the DFI of the “good quality” group after Uselect selection as it already had sperms of good quality (P = 0.83). In the “DFI moderate” subgroup, 5 out of 14 (35.7%) samples showed improvement in DFI from the moderate to good DFI class, and another 8 samples (57.1%) showed improved DFI. In the poor DFI subgroup, improvement in DFI was highly significant (P <0.001), with a 44.9% reduction after USelect separation. In the poor DFI class, except for two, all the samples showed improvement in the DFI classification. The USelect technique was applied to 22 samples. Results showed that 77.3% of the samples improved to a moderate DFI, and 9.1% achieved a good DFI. Figure 5c visually represents these outcomes, indicating the effectiveness of the USelect technique in enhancing the selection of sperms with better DFI. The HA rebinding test was performed on the sperms selected by all three methods - Uselect, DG, and SU. USelect selected sperms have shown rebinding of 63.6% of sperm to HA while the same from the other two methods were 44.8% and 19.6% respectively (Figure 5d). This ensures that these sperms can efficiently bind to the HA of the oocyte.

**Figure 5:**
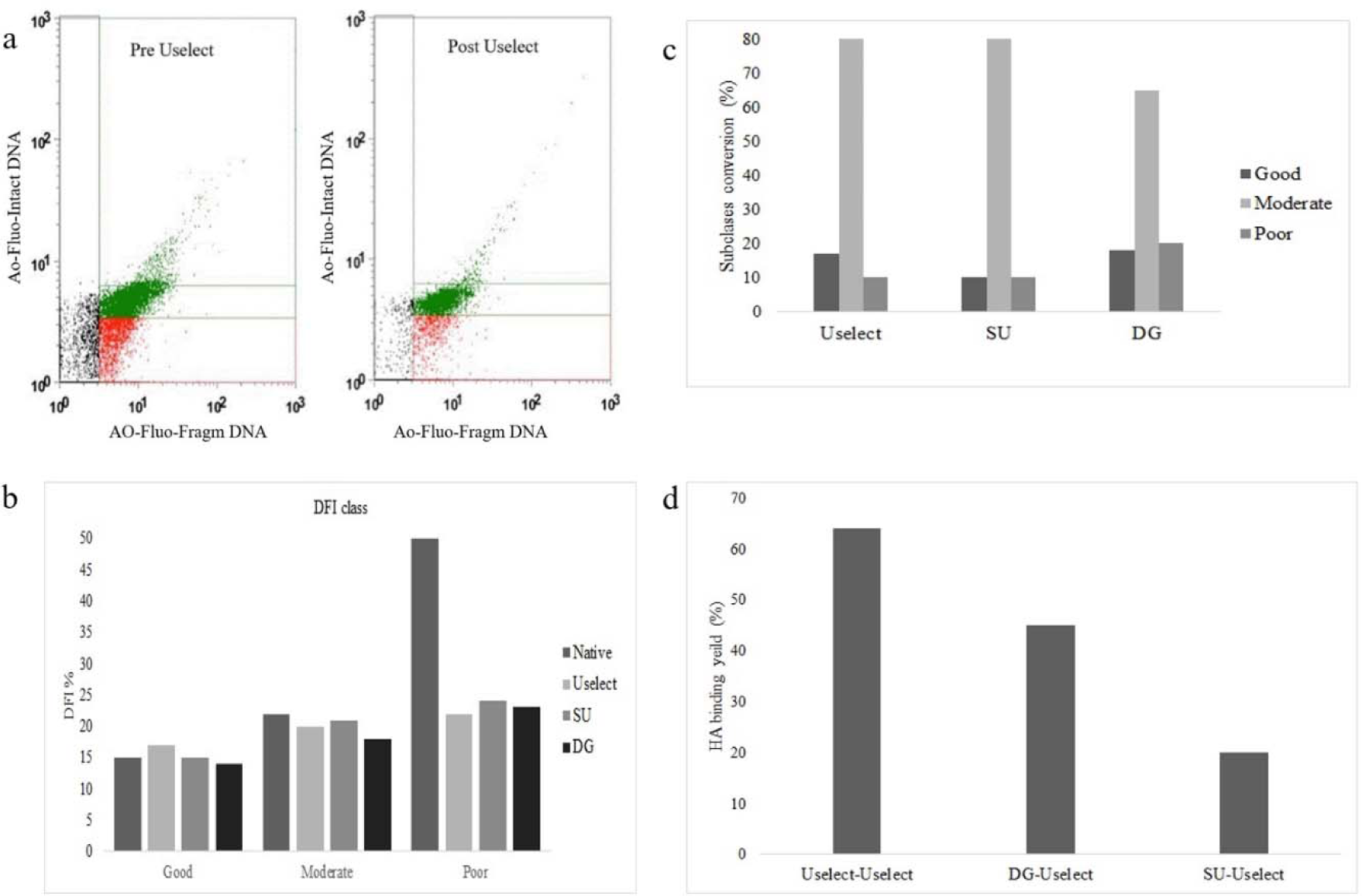
Comparison of USelect, DG, and SU techniques for isolation of viable and mature sperms. Comparison of USelect, DG, and SU techniques for isolation of viable and mature sperms. (a) Flow cytometric scatter plot of sperm population in pre and post Uselect selection. Green dots-sperms with Intact DNA, red dots-sperms with fragmented DNA, and black dots-non-sperm debris. (b) Sub-group analysis of DNA integrity of native sperms and sperms processed by USelect, DG and SU techniques. Good (DFI ≤ 15%) n=2, moderate (DFI 15-30%) n=14, poor (DFI >30%) n=22. Mean DFI of sperms isolated using USelect technique was significantly lower P<0.001 in moderate and poor DFI classes when compared to native sperms. (c) Improvement in DFI class of poor DFI sperms after processing by USelect, SU and DG methods. (d) HA binding efficiency of sperms isolated using USelect, SU and DG techniques. Sperms from poor subclass were first isolated using each technique followed by incubation with HA beads and re-isolation using USelect technique (n=6).

**Table 1:** Semen analysis characteristics, viability, and DNA integrity of unprocessed (native) and sperms processed by different separation techniques.

|  | Native | Uselect | DG | SU |
| --- | --- | --- | --- | --- |
| **Sperm count** (millions / mL) |  |  |  |  |
| Mean ± SD | 73.15 ± 23.14*** | 26.00 ± 22.38 | 49.21 ± 31.76<sup>\$</sup> | 24.17 ± 24.45 |
| Range | 28 - 152 | 2 – 100 | 2 - 120 | 1 - 100 |
| **Motility** (%) |  |  |  |  |
| Mean ± SD | 43.05 ± 8.87 | 34.05 ± 23.18 | 59.45 ± 26.55** | 40.45 ± 25.70 |
| Range | 20 - 60 | 0 – 90 | 15 - 90 | 5 - 90 |
| **DNA Fragmentation Index** (%) |  |  |  |  |
| Mean ± SD | 36.99 ± 16.65*** | 20.94 ± 9.62 | 18.76 ± 12.11 | 22.29 ± 12.79 |
| Range | 9.53 - 77.97 | 6.21 - 61.14 | 4.21 - 58.87 | 4.95 - 53.18 |

\*\*\* P<0.001, \*\* P<0.01 One-way ANOVA using Tukey's multiple comparison test. <sup>\$</sup>MAGNETIC vs. DG P<0.001, <sup>§</sup>Student's *t*-test P=0.99 (Morphology), P=0.79 (HOS)

To further understand the benefits and performance of the USelect technique beyond the *in vitro* study, a clinical study was conducted to compare clinical pregnancy outcomes between USelect and the commonly used DG technique. Subjects for the clinical study were stratified through the randomized selection of patients that included 50 women in the DG group and 50 women in the USelect group. Enrolling the first 50 women in the Uselect group and the next 50 in the DG group ensures a systematic and fair distribution of participants, aiming for a representative sample and minimizing potential biases. This approach is for randomization control, establishes comparability at baseline, and enhances the validity of subsequent group comparisons. Biochemical pregnancy was confirmed by testing β-HCG levels at the end of three weeks. The positive pregnancy rate was higher in the USelect group when compared to the DG group [(32% (n =16 positive out of 50) vs. 18% (n = 9 positive out of 50)]. In the USelect group, 13 out of 16 positive pregnancies occurred in the first IUI cycle (81% of total positive pregnancies and 26% of total samples n=50). Under the USelect group, four patients were considered for the second IUI cycle by the treating clinician, out of which 3 achieved positive pregnancy (19% of total pregnancies and 6% of the total sample n=50). In the DG group, 7 out of 9 pregnancies occurred on the first IUI cycle (78% of total positive pregnancies and 14% of total samples). In the DG group, 2^nd^ IUI was performed in a total of 9 subjects, out of which 2 pregnancies occurred (22% of total positive pregnancies and 4% of the total sample). Samples under two or more IUI cycles have shown a 75% positive rate under USelect compared to 22 % in DG (Figure 6A).

**Fig. 6:**
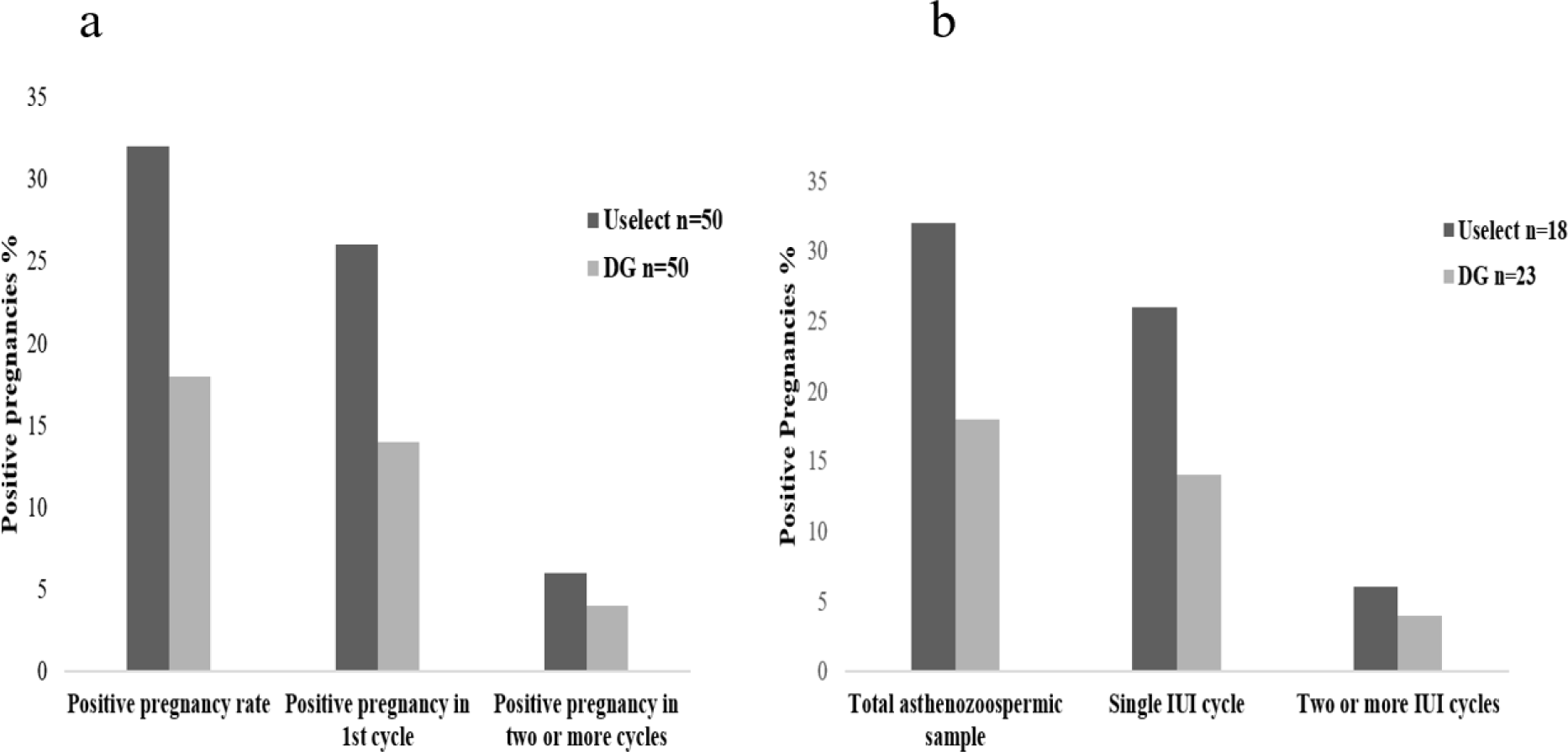
Comparison of USelect versus DG pregnancy rates between (a) total number of samples, in single and 267 two or more IUI cycles in complete sample size, (n=50under both USelect and DG) (b) total number of samples, 268 in single and two or more IUI cycles in mild asthenozoospermic samples (USelect n=18, DG n=23).

Under USelect, group 18 out of 50 samples were mild asthenozoospermic (15 to 38% motility), however, 8 out of these 18 were biochemically positive, which gives rise to 44 % positive cases (n=18). Among these 8 positives, 6 attained successes in a single IUI cycle, which gives rise to a 33% success rate. Among four samples which are taken for the second IUI cycle, two were mild asthenozoospermic and both attained positive biochemically (11%). In the DG group 23 out of 50 samples were mild asthenozoospermic (15 to 38% motility), however, 4 out of these 23 were biochemically positive, which gives rise to 17% positive cases (n=23). Among these, 3 attained successes in a single IUI cycle, which gives rise to a 13% success rate. Among 9 samples that are taken for the second IUI cycle, three are mild asthenozoospermic and one is positive biochemically (4%) in the second IUI cycle (Figure 6B).

## DISCUSSION

Sperm selection plays a vital role in ART treatment, as it ensures the selection of the finest quality sperm for fertilization, thereby increasing the likelihood of a favorable outcome, which is not achievable through conventional methods [22]. Among the essential techniques for sperm separation, DG, and SU stand out as significant and widely used approaches [14,15]. Ongoing research aims to comprehensively assess their advantages and limitations, working towards the refinement of sperm selection strategies. However, these methods have limitations. While DG causes diminished sperm recovery due to centrifugation-related damage, SU’s effectiveness can fluctuate based on factors such as motility and time-dependent recovery rate [23,24]. We have aimed to overcome some of these challenges while designing the USelect method.

It is well known that HA has a role in the natural selection of sperm. In normal reproduction, HA present in the extracellular matrix of the cumulus oophorus complex surrounding the oocyte act as the physiological selector of sperm. As a result, only sperms with acrosomal maturity can bind to the oocyte layer and initiate the cascade of reactions that lead to fertilization. Therefore, the potential of HA-based selection may be better understood when tested in IUI as fertilization occurs *in vivo*. However, because the PICSI plate-based sperm selection yields only a few sperm that bind to HA, this method cannot be applied in IUI, which needs millions of sperm to achieve a successful pregnancy. The USelect technology aims to overcome these challenges by establishing a robust protocol for the extraction of millions of HA-binding sperms within 30-40 min. The developed technique is simple, requiring only basic lab equipment that is widely available along with a training protocol that is easy to learn.

In a 2-step process, washed semen samples are first incubated with HA-coated magnetic beads leading to the binding and separation of the desired sperm followed by a wash step with a buffer that releases the HA-bound sperm for use in an IVF procedure. The protocol for isolating mature and competent sperm has been optimized for conditions such as the concentration and volume of HA-bound magnetic beads, and the pH and hyaluronidase concentration for the sperm release buffer. The optimal protocol uses 60µg of beads per sample, with the sperm release step being conducted at pH 7.0 using 100 U hyaluronidase concentrations (Supplementary Figure 1).

The study further visually investigated the binding abilities of sperm to beads coated with HA protein. Results showed that mature sperm have multiple HA-binding proteins on their surface, which enables a single sperm to bind to multiple beads. When semen samples were incubated with the HA-coated magnetic beads, motile sperm were seen attached to the beads, with some carrying one or more beads attached to the surface of their sperm head (Figure 2). In the acute toxicity assay, the animals showed neither clinical signs of toxicity nor mortality up to 48 hours after the IV administration even when reagent beads at high concentrations used in the process were injected, demonstrating their safety. In practice, no beads or very small concentrations of beads are expected to be seen as carry over from the process. Both motile and a small fraction of immotile sperm were selected by the technique showing that motility and HA-binding are not necessarily correlated with each other (Data not shown).

In our current research, we undertook a similar comparison of three sperm selection methods: Uselect, DG, and SU. Our analysis specifically focused on comparing the DFI. Notably, the results revealed a significant enhancement in DFI values for samples processed using the Uselect method compared to DG and SU methods. Importantly, we found that the USelect selected good quality sperms with significantly reduced DNA fragmentation and high DFI. The increased percentage of sperm DNA fragmentation is detrimental to achieving and sustaining pregnancies and is associated with recurrent pregnancy loss. This could be avoided when Uselect is used. A four-fold increase in miscarriage risk has been reported in IVF and ICSI data regarding cases with elevated DNA fragmentation [25,26]. In this regard, the present USelect technique is an easy, convenient, and cost-effective method to select competent sperms with intact acrosomal status and better DNA integrity compared to native, unprocessed sperms. The reduction in DFI for poor and moderate samples was equivalent to that found with SU and DG techniques (Table 1, Figure 5c).

We conducted tests to assess the ability of sperms isolated using three different techniques to bind with HA. Mature sperms need to exhibit HA binding ability, considering their unreacted acrosomal status. Our findings revealed that sperms isolated through SU and DG methods had 1.6- and 3 times lower HA binding efficiency, respectively, compared to those isolated through the USelect technique from the same sample. Furthermore, the capacity of sperms to re-bind with HA is crucial as it indicates that USelect-selected sperms retain their ability to bind with the oocyte.

After the findings of the *in vitro* study were completed, the USelect technique was then extended to clinical usage after the standardization of the protocol and the sample study described above. The study compared 50 patients who were subjected to IUI (USelect) to 50 other patients who were subjected to IUI (DG). Overall, the USelect cases resulted in 16 (32%) biochemical pregnancies while the DG cases resulted in 5 (10%) biochemical pregnancies when compared across 50 patients in each arm.

Specifically, 26% (n=13) of all samples extracted using USelect (n=50) resulted in pregnancy in the first cycle itself, compared to only 6% (n=3) using the DG method (Figure 6a). The study revealed that the success rate of pregnancies in the first IUI cycle was notably higher in samples that were extracted using the USelect method. The study was non-interventional and hence, the decision to take patients for more than one IUI cycle was purely based on clinical findings and laboratory investigations, and not on the group under which samples were stratified. Clinicians decided to take a total of 13 patients to the second or more than two IUI cycles. Among these, 4 were from the USelect group and 9 were from the DG group. In the second cycle, 3 out of those 4 patients from the USelect group were biochemically positive while 2 out of 9 patients were positive in the DG group, (Figure 6a). In total, the samples processed with USelect gave 32% biochemically positive results in a single IUI attempt, even though the samples were mild asthenozoospermic (n=18). Biochemical pregnancies were lower by 50% (n=9) using samples processed with DG (Figure 6b). The data suggests that USelect outperforms DG and shows almost twice the positive pregnancy rate while performing an IUI. Thus far, HA-based sperm selection such as PICSI has been used in advanced ART techniques with mixed results. While several studies reported improvements in DNA integrity and higher pregnancy success rates [20,21], others have shown no difference in the rates of live births or clinical pregnancies between PICSI and ICSI although outcomes in older women were improved [27,28]. However, since HA-bound sperm have not been available for use in IUI, there is no direct comparison available in the literature. We believe this study represents the first attempt to use HA-bound sperm for IUIs.

## CONCLUSION

In conclusion, the USelect technique is a simple and uncomplicated procedure, does not involve repeated centrifugation like conventional SU and DG methods, and yields a sufficiently high count of mature sperms for use in IUI. The USelect selected sperms showed significant DFI and HA binding capacity that led to an increased success rate in IUI among 50 study subjects. Based on the findings of this work, we propose that the USelect technique can be used for the HA-based selection of acrosome-matured sperm and is suitable for application in IUI. The study’s limitation lies in the suboptimal sperm motility observed after the SU and DG selection. It is crucial to note, though, that the research specifically focused on male partners within infertile couples, so poor motility is to be expected in many of these samples. However, multicentric and multiethnic randomized clinical trials with higher patient sample sizes are required to generate further insights and confirmation of the advantages of USelect. Also, establishing live birth rates in comparison to existing methodologies such as DG and SU will be essential to confirm the success of the technique.

## PATENT

Year of publication 2022, “Method for extracting viable sperms from a semen sample”, Patent number 396145, date of filing: 09/09/2016, and issued date of publication: 05/04/2022

## AUTHOR CONTRIBUTION

DD and VS conceptualized and designed the study, MS performed the assay and clinical validation. MS and CRN analyzed the data. MS and CRN wrote the manuscript. All authors have read and approved the final manuscript.

## ACKNOWLEDGMENTS

We acknowledge the contributions of the Doctors and the staff members of Manipal-Ankur Andrology and Reproductive Services and Achira Labs Pvt Ltd.

## DATA AVAILABILITY

The datasets used and/or analysed during the current study available from the corresponding author on reasonable request.

